# Personalized allele-specific CRISPR-Cas9 strategies for myofibrillar myopathy 6

**DOI:** 10.1101/2024.02.03.24302252

**Authors:** Jun Wan Shin, Kyung-Hee Kim, Yukyeong Lee, Doo Eun Choi, Jong-Min Lee

## Abstract

Myofibrillar myopathy 6 (MFM6) is a rare childhood-onset myopathy characterized by myofibrillar disintegration, muscle weakness, and cardiomyopathy. The genetic cause of MFM6 is p.Pro209Leu mutation (rs121918312-T) in the *BAG3* gene, which generates the disease outcomes in a dominant fashion. Since the consequences of the *BAG3* mutation are strong and rapidly progressing, most MFM6 patients are due to *de novo* mutation. There are no effective treatments for MFM6 despite its well-known genetic cause. Given p.Pro209Leu mutation is dominant, regenerative medicine approaches employing orthologous stem cells in which mutant *BAG3* is inactivated offer a promising avenue. Here, we developed personalized allele-specific CRISPR-Cas9 strategies capitalizing on PAM-altering SNP and PAM-proximal SNP. In order to identify the disease chromosome carrying the *de novo* mutation in our two affected individuals, haplotype phasing through cloning-sequencing was performed. Based on the sequence differences between mutant and normal *BAG3*, we developed personalized allele-specific CRISPR-Cas9 strategies to selectively inactivate the mutant allele 1) by preventing the transcription of the mutant *BAG3* and 2) by inducing nonsense-mediated decay (NMD) of mutant *BAG3* mRNA. Subsequent experimental validation in patient-derived induced pluripotent stem cell (iPSC) lines showed complete allele specificities of our CRISPR-Cas9 strategies and molecular consequences attributable to inactivated mutant *BAG3*. In addition, mutant allele-specific CRISPR-Cas9 targeting did not alter the characteristics of iPSC or the capacity to differentiate into cardiomyocytes. Together, our data demonstrate the feasibility and potential of personalized allele-specific CRISPR-Cas9 approaches to selectively inactivate the mutant *BAG3* to generate cell resources for regenerative medicine approaches for MFM6.

## Introduction

Myofibrillar myopathy 6 (MFM6; MIM 612954) is a childhood-onset myofibrillar myopathy [1]. Most MFM6 subjects are severely affected as the clinical symptoms of MFM6 begin in the first decade and progress rapidly, leading to proximal muscle weakness, respiratory insufficiency, cardiomyopathy, and skeletal deformities [2-5]. The genetic cause of MFM6 is a mutation changing the proline at the 209th amino acid of BCL2-associated athanogene 3 (BAG3) protein to leucine (p.Pro209Leu; rs121918312) [4, 5]. Since the disease is dominant and severe, most MFM6 cases result either from *de novo* mutation or from inheritance of the mutation from a parent who is a genetic mosaic for the mutation [4, 6-8]. Although certain small molecule drugs showed improvements in pre-clinical models [9-12], there are no effective treatments for MFM6. Therefore, we reasoned that advancements in genome engineering may provide alternative routes for effective treatment strategies for MFM6, given its well-known genetic underpinning. Understanding how p.Pro209Leu mutation produces MFM6 became critical to select appropriate therapeutic gene editing strategies for this devastating disease.

Since the p.Pro209Leu mutation in BAG3 is dominant [5, 13] and heterozygous *Bag3* knock-out does not produce muscle phenotypes in mice [14, 15], it is hypothesized that MFM6 is generated through gain-of-function mechanisms. In support, reduced BAG3 levels due to nonsense mutation, frameshift mutation, or genomic deletion produced dilated cardiomyopathy (DCM) [16-19], which is less severe compared to MFM6. However, gain-of-function leading to deficiency also has been proposed [12, 20], suggesting that BAG3 protein with p.Pro209Leu mutation acts as a dominant negative. Considering these findings, genetic correction of the mutation by converting T to C at rs121918312 using CRISPR-Cas9 or base editing appeared to be the ideal therapeutic gene editing strategy. However, 1) a robust CRISPR PAM site (i.e., NGG) for optimal conversion of A to G at rs121918312 is lacking, and 2) base editing utilizing other PAM sequences has the risk of converting neighboring bystander nucleotides due to its somewhat wide conversion windows [21-23]. Prime editing may suffer from safety concerns as enhanced editing may require the suppression of endogenous DNA repair genes [24-26].

Given 1) technical difficulties in correcting the disease-causing mutation, 2) p.Pro209Leu mutation is dominant, and 3) heterozygous knock-out does not produce MFM6, selectively inactivating the mutant *BAG3* may be a viable alternative approach for MFM6. Recently, we developed allele-specific CRISPR-Cas9 strategies capitalizing on the genetic variations that generate or eliminate the protospacer adjacent motif (PAM) sequence for CRISPR-Cas9 [27-29]. Here, we extended our allele-specific CRISPR-Cas9 strategies to develop personalized mutant-selective gene editing approaches for our two MFM6 subjects carrying the p.Pro209Leu mutation. Subsequently, we applied novel allele-specific CRISPR-Cas9 strategies on patient-derived cells, generating autologous induced pluripotent stem cells (iPSCs) with mutant *BAG3* allele inactivated, representing cell resources with direct applicability in the realm of regenerative medicine.

## Results

### Allele-specific CRISPR-Cas9 strategies using PAM-altering SNPs or PAM-proximal SNPs

We previously developed allele-specific CRISPR-Cas9 strategies using DNA variations that generate or eliminate the CRISPR-CAS9 PAM sequence (i.e., PAM-altering SNP; S. Fig. 1A), and demonstrated their high levels of allele specificities [27-29]. In addition, genetic variations that introduce mismatches near the PAM site can be used to edit the gene in an allele-selective manner (i.e., PAM-proximal SNP; S. Fig. 1B) [30]. When utilizing PAM-altering SNP (PAS) or PAM-proximal SNP (PPS), the dominant mutation can be inactivated by two different modes of CRISPR-Cas9, such as Transcription-Prevention CRISPR (TP-CRISPR) and Nonsense-Mediated Decay CRISPR (NMD-CRISPR). In the TP-CRISPR approach, two guide RNAs (gRNAs), including at least one allele-specific gRNA, are simultaneously used to excise a genomic region harboring the transcription start site (TSS) of the mutant allele. Allele-specific genomic excision disallows the transcription of the mutant gene, preventing the generation of mutant RNA or protein (S. Fig. 2A) [29]. In contrast, allele-specific NMD-CRISPR relies on one allele-selective gRNA that targets the coding region of the mutant allele. Therefore, allele-specific NMD-CRISPR generates small indels on the mutant gene, leading to the degradation of mutant mRNA via NMD (S. Fig. 2B).

TP-CRISPR strategies do not edit the disease-causing mutations directly; instead, they are designed to target the haplotype that carries the disease-causing mutation. Since there are numerous PAS suitable for TP-CRISPR [31], this approach can be applied to many genes [29]. However, the use of two gRNAs may decrease the editing efficiencies and increase off-targeting. By contrast, NMD-CRISPR may show increased editing efficiencies and decreased off-targeting. However, if nonsense-mediated decay is incomplete or fragment protein is produced, NMD-CRISPR may generate incomplete inactivation or unexpected outcomes, respectively. In addition, allele-specific NMD-CRISPR may be limited by the availability of PAS and PPS due to their relative scarcity in the coding sequences, representing one of the shortcomings of NMD-CRISPR.

### Haplotype phasing to determine the chromosomes with *do novo* p.Pro209Leu mutations

Developing allele-specific TP-CRISPR and NMD-CRISPR strategies for our MFM6 subjects required the identification of PAS and PPS as the first step. Our two independent probands (subjects #3 and #6) carry *de novo* mutation as none of the parents has rs121918312-T allele (Fig. 1A). Therefore, it was not immediately clear whether the disease-causing allele is on the paternal or maternal chromosome, posing challenges in developing allele-specific CRISPR-Cas9 strategies. To facilitate the identification of the allele-specific target sites, we determined the phase of the mutant allele (rs121918312-T) in our two affected individuals. Briefly, we utilized a neighboring SNP to determine whether *de novo* mutations have occurred on the paternal or maternal chromosome. Since both probands (subjects #3 and #6) are heterozygous at rs196330 (located approximately 4 Kb downstream), and parents carry different genotypes at the same locus, we took advantage of this SNP to infer haplotype phase (S. Fig. 3). Sequencing of cloned DNA regions from subject #3 (affected child in family A) revealed that the disease-causing mutation in this proband occurred on the paternal chromosome (Fig. 1B). In contrast, subject #6 (affected child in family B) was explained by a *de novo* mutation on the maternal chromosome (Fig. 1C). Importantly, these data indicated that allele-specific CRISPR-Cas9 targeting the paternal and maternal allele would inactivate the disease-causing mutation in subjects #3 and #6, respectively.

**Figure 1.**
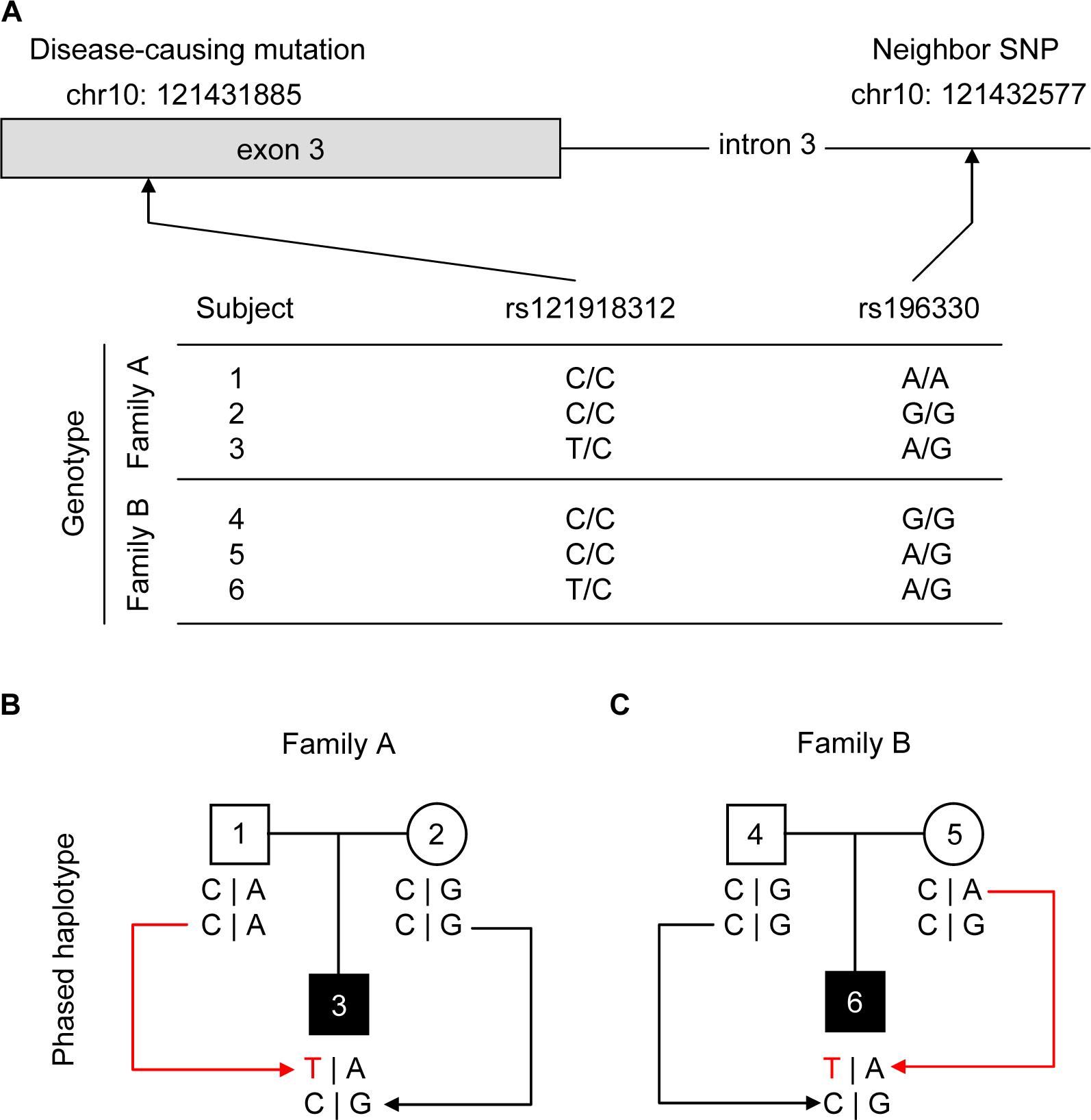
Inferring the haplotype phase of the *de novo* mutations responsible for MFM6 in the study subjects. A) To determine whether the *de novo* mutation (rs121918312-T in exon 3) occurred on the paternal or maternal chromosomes, we used a neighbor SNP, which differed between the father and the mother. Unaffected parents carry different alleles at an intronic SNP rs196330, allowing the inference of the haplotype that carries the disease-causing mutation in the probands. B) Genotypes of the disease-causing mutation (rs121918312) and a neighbor SNP (rs196330) are displayed for study participants, including two probands (subjects #3 and 6; filled black). Red lines and letters represent *de novo* mutation and disease-causing allele, respectively.

### Personalized allele-specific CRISPR-Cas9 strategies for MFM6

Subsequently, we performed a series of sequencing analyses for each trio to identify genetic variations that permit mutant-specific TP-CRISPR and NMD-CRISPR. Since PAS approaches demonstrated high levels of allele specificity, we focused on DNA variants that alter the NGG PAM sequence [27-29]. For TP-CRISPR strategies for subject #3, we could not identify allele-specific PAS upstream of the TSS due to many homopolymers around the candidate sites. However, we found that rs3847489, located at the first intron, produces NGG PAM site selectively on the paternal chromosome (i.e., mutant *BAG3*). Therefore, we designed an allele-specific TP-CRISPR strategy for subject #3 using a combination of one non-allele-specific (chr10:121410519) and one allele-specific gRNAs (Fig. 2A). This TP-CRISPR strategy designed specifically for subject #3 was expected to excise approximately 6.8 Kb genomic region from the paternal chromosome, resulting in the prevention of the transcription of the mutant *BAG3*. In contrast, an allele-specific CRISPR-Cas9 strategy targeting using two mutant-specific sites was feasible for subject #6 as the maternal chromosome (i.e., mutant allele) of this subject carries the NGG PAM site generated by rs196290 (upstream of TSS) and rs11199061 (downstream of TSS). The TP-CRISPR strategy was expected to result in genomic deletion of 6.7 Kb in the mutant *BAG3 of* subject #6, preventing the production of the mutant *BAG3* (Fig. 2B).

**Figure 2.**
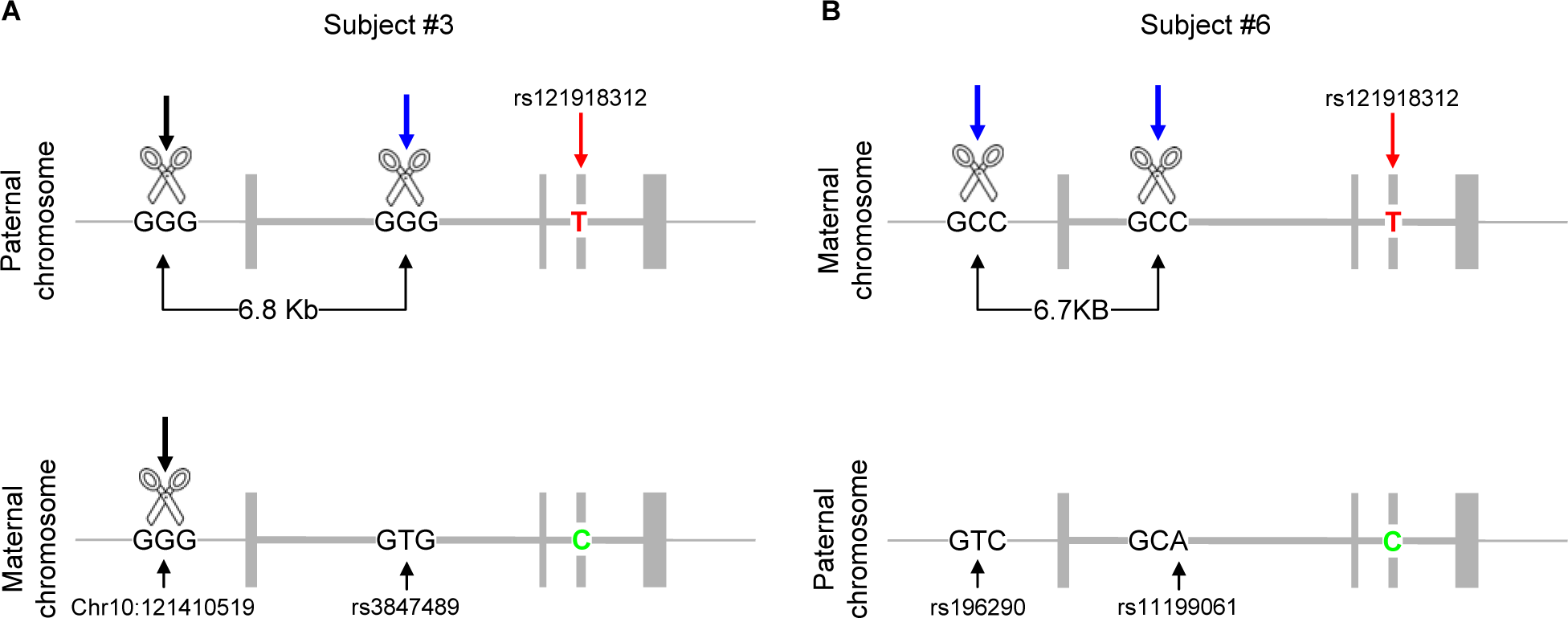
Individualized allele-specific TP-CRISPR strategies for study participants using PAS. A) An allele-specific TP-CRISPR strategy designed for subject #3 is summarized. This strategy utilized one non-allele-specific gRNA targeting an NGG PAM site at Chr10:121410519 and one allele-specific gRNA targeting an NGG PAM site generated by rs3847489. This strategy was expected to excise 6.8KB DNA (involving the TSS) from the mutant *BAG3* allele. B) An allele-specific TP-CRISPR strategy for subject #6 utilizes two allele-specific NGG PAM sites generated by rs196290 and rs11199061. The TP-CRISPR strategy for subject #6 was expected to excise 6.7KB DNA from the mutant allele involving the TSS. Black and blue arrows represent non-allele-specific and allele-specific editing, respectively. DNA alleles in green and red indicate mutant and normal alleles at rs121918312.

Unfortunately, coding sequences of neither proband carry mutant *BAG3*-specific NGG PAM sites, not permitting CRISPR-9 strategies utilizing PAS. However, the disease-causing variant (rs121918312) is an actual PAM-proximal SNP (4th nucleotide from the PAM), permitting allele-specific NMD-CRISPR strategy for all MFM6 (Fig. 3).

**Figure 3.**
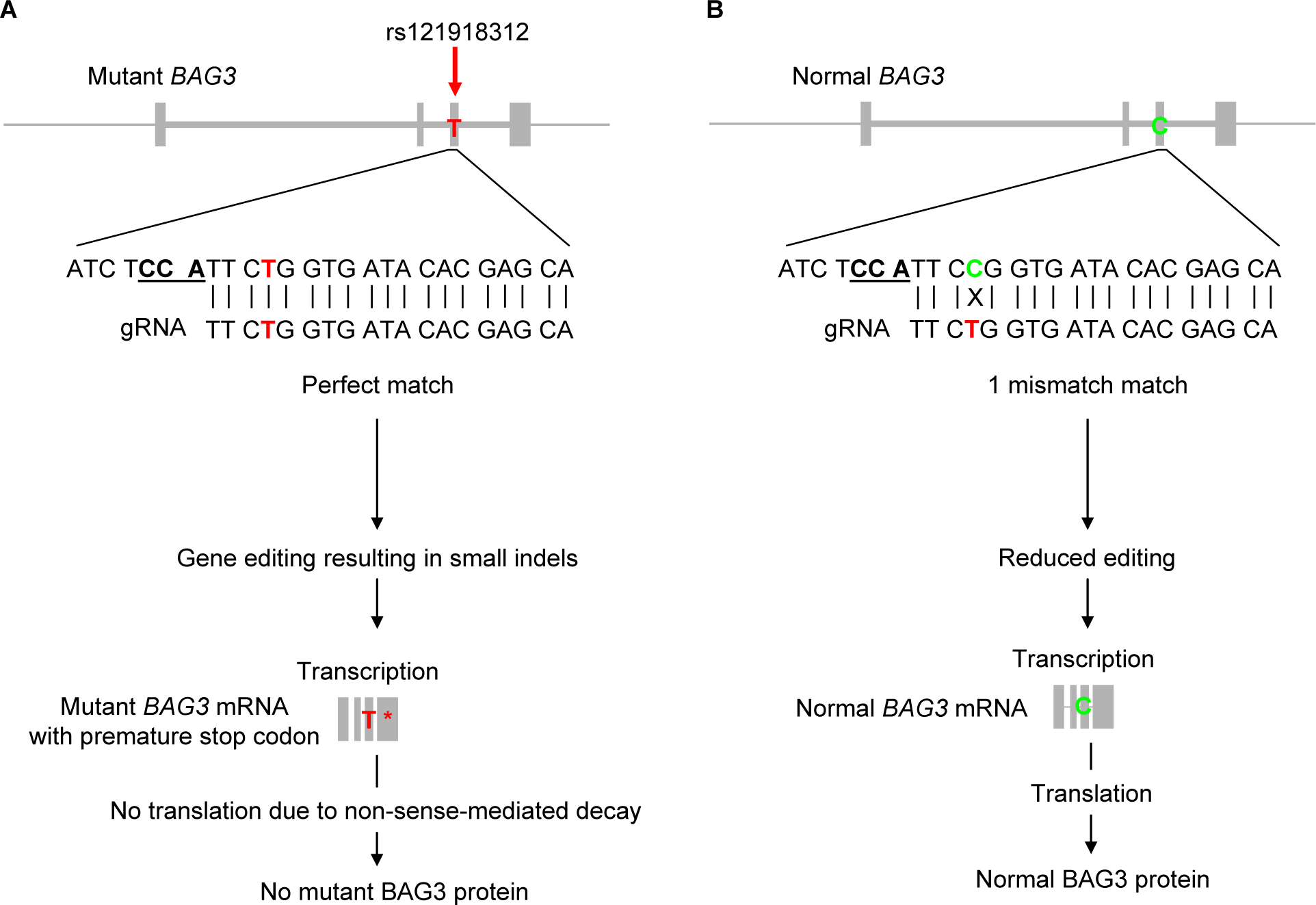
Allele-specific NMD-CRISPR strategy for MFM6 using the disease-causing mutation as the PSS. Since a nearby NGG PAM site flanks the disease-causing mutation (rs121918312), we designed a CRISPR-Cas9 gRNA using the rs121918312 as the PPS to achieve allele-specific NMD-CRISPR. In this scenario, a gRNA designed to target the mutant allele (19 nucleotide gRNA is displayed) will interact with the mutant allele (A) without mismatches. However, the same gRNA is expected to interact with normal BAG3 with one mismatch (B), potentially leading to mutant-specific NMD.

### Selectivity of allele-specific CRISPR-Cas9 strategies

We then experimentally determined the levels of allele specificity of personalized allele-specific CRISPR-Cas9 strategies in patient-derived iPSC lines. Patient-derived cells were treated with personalized allele-specific TP- and NMD-CRISPR strategies. For TP-CRISPR strategies, we amplified the DNA region encompassing the predicted genomic deletion and sequenced using a MiSeq platform to determine the proportion of edited mutant and normal *BAG3* alleles. Due to the large genomic deletion expected from our TP-CRISPR approaches, evaluating the editing efficiencies of TP-CRISPR approaches was technically challenging. Nevertheless, if focusing on the edited alleles that were PCR amplified, sequenced, and quality control (QC)-passed, all sequence reads (representing targeted alleles) contained rs121918312-T allele, indicating that our allele-specific TP-CRISPR strategies for subjects #3 and #6 did not target the normal *BAG3* at all (Table 1). It is important to note that the TP-CRISPR strategy for subject #3 using a combination of one non-allele-specific and one allele-specific gRNAs also resulted in a complete mutant allele-specificity, suggesting that one allele-specific gRNA may be sufficient to achieve high levels of allele-specificity in the TP-CRISPR approaches based on PAS (Table 1).

**Table 1.**
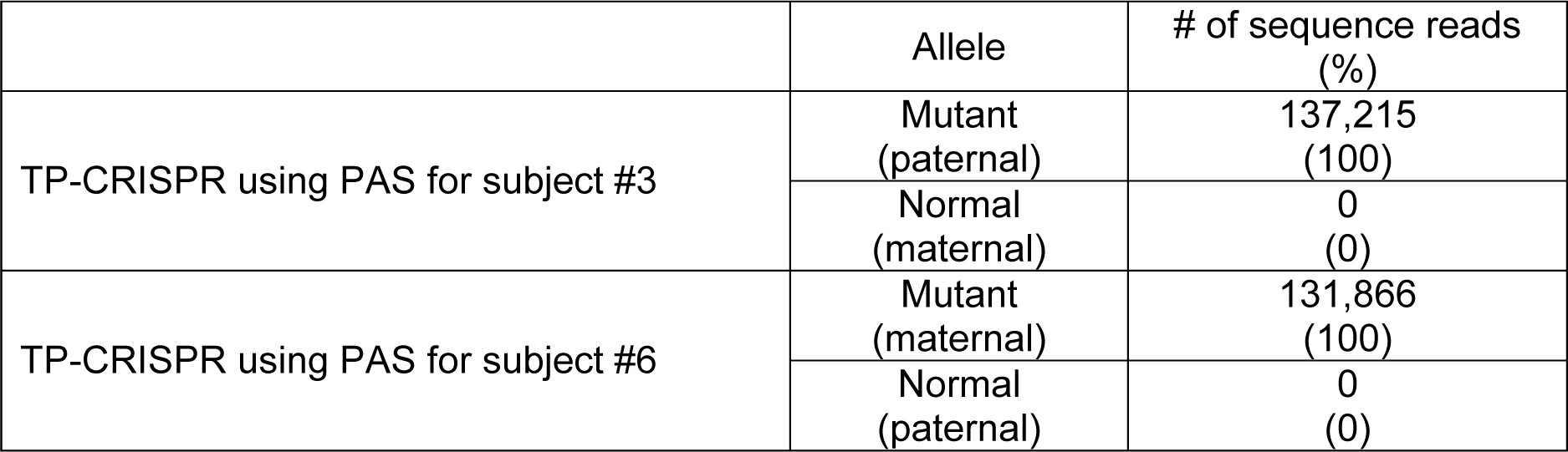
Allele specificity of personalized TP-CRISPR strategies. Patient-derived iPSC lines were treated with personalized allele-specific TP-CRISPR strategies. Subsequently, DNA samples were subjected to PCR to amplify the genomic region with deletion. The identity of alleles was determined based on the alleles of PAS.

|  | Allele | # of sequence reads<br>(%) |
| --- | --- | --- |
| TP-CRISPR using PAS for subject #3 | Mutant<br>(paternal) | 137,215<br>(100) |
|  | Normal<br>(maternal) | 0<br>(0) |
| TP-CRISPR using PAS for subject #6 | Mutant<br>(maternal) | 131,866<br>(100) |
|  | Normal<br>(paternal) | 0<br>(0) |

Since our allele-specific NMD-CRISPR was predicted to produce small indels, determining the editing efficiency was more feasible compared to TP-CRISPR. As previously observed [27, 28, 32-34], editing efficiencies of CRISPR-Cas9 strategies in iPSC were modest. Nevertheless, our allele-specific NMD-CRISPR strategy using a PPS showed complete allele specificities (Table 2). For example, our NMD-CRISPR strategy produced small indels in the 4.6% and 4.1% of mutant *BAG3* in subjects #3 and #6, respectively, while the same treatment did not target the normal counterpart (Table 2). Together, these data suggested that our allele-specific CRISPR-Cas9 approaches are highly allele-specific, illustrating the feasibility of individualized CRISPR-Cas9 approaches to inactivate the mutant *BAG3* gene selectively.

**Table 2.** Editing efficiency and allele specificity of allele-specific CRISPR-Cas9. Patient-derived iPSC lines were treated with the allele-specific NMD-CRISPR strategy. Subsequently, DNA samples were subjected to PCR to amplify the region harboring rs121918312, and then MiSeq sequencing analysis was performed. The identity of alleles was inferred based on the alleles of rs121918312.

|  | Allele | Modification | Subject #3 (%) | Subject #6 (%) |
| --- | --- | --- | --- | --- |
| NMD-CRISPR using PAS | Mutant | Modified | 4.6 | 4.1 |
|  |  | Unmodified | 41.8 | 43.4 |
|  | Normal | Modified | 0 | 0 |
|  |  | Unmodified | 53.6 | 52.5 |

### Molecular outcomes of allele-specific CRISPR-Cas9 strategies for MFM6

Since our allele-specific NMD-CRISPR strategy targeting rs121918312 as the PPS was highly allele-specific and widely applicable, we further investigated the molecular consequences of the mutant-specific NMD-CRISPR strategy. To establish targeted clonal lines, we treated patient-derived iPSCs with allele-specific NMD-CRISPR (Fig. 3). For each study subject, we developed four independent targeted iPSC clonal lines. Analysis of genomic DNA using a MiSeq platform determined a frameshift mutation the mutant *BAG3* in each clonal line (S. Fig. 4). In addition, MiSeq analysis of RNA from the targeted clonal lines confirmed the same frameshift genome modifications that were detected in the DNA (Fig. 5A). It is important to note that the levels of mutant *BAG3* mRNA were not zero (Fig. 4A) even though each clonal line carries a frameshift mutation that was expected to induce NMD (Fig. 5A). This might be due to the possibility that newly synthesized mutant *BAG3* mRNA has not been subjected to NMD yet, which was observed previously [28]. Still, we predict that the amount of mutant BAG3 protein would be very low, considering newly generated premature stop codons in each clonal line (S. Fig. 5A). In support, immunoblot assays showed significantly decreased total BAG3 protein levels (Fig. 4B). Given only mutant *BAG3* was edited, reduced BAG3 protein levels were attributable to decreased mutant protein, supporting its allele specificity.

**Figure 4.**
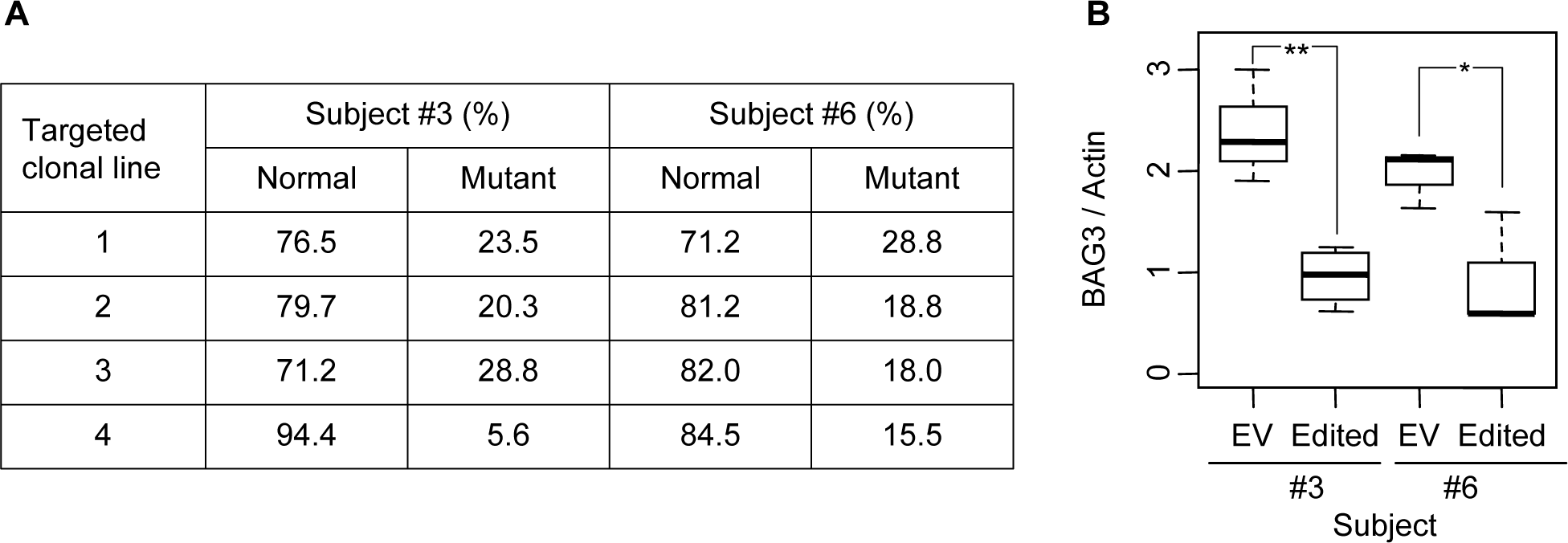
Molecular consequences of allele-specific NMD-CRISPR. A) MiSeq analysis was performed on mRNA samples from targeted clonal lines from subjects #3 and #6 to evaluate the levels of *BAG3* mRNA containing normal and mutant alleles at rs121918312. B) Immunoblot analysis results representing normalized total BAG3 protein are displayed. Three and four independent clonal lines were analyzed for each proband. *, p-value < 0.05 and **, p-value < 0.01. EV, empty vector.

**Figure 5.**
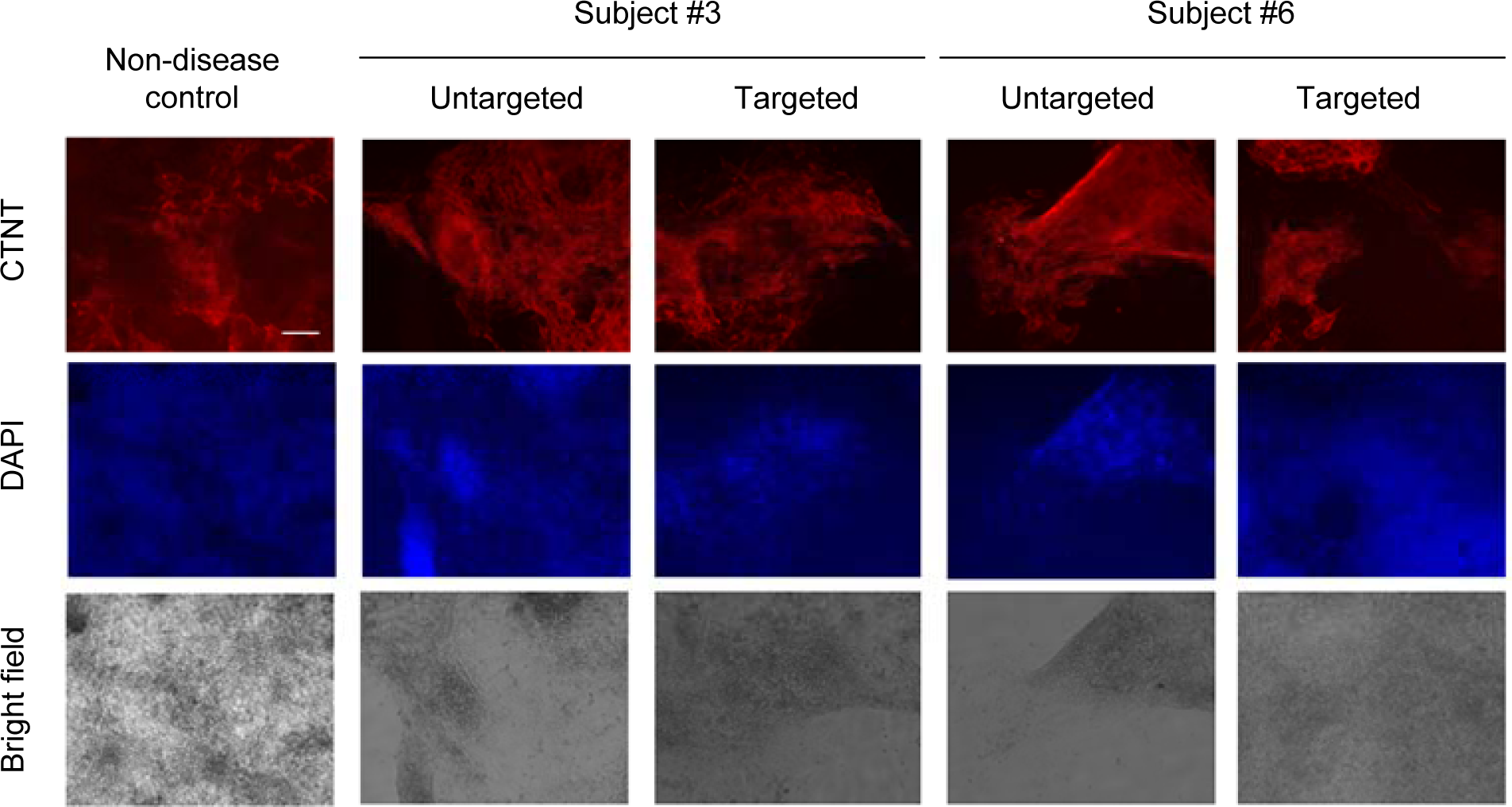
Differentiation into cardiomyocyte from MFM6 patient-derived iPSCs. Immunocytochemistry of CTNT was performed for non-disease control, untargeted, and targeted clonal lines from subject #3 and subject #6 to evaluate the capacity of differentiation into a cardiomyocyte. DAPI staining shows nuclei. The scale bar in the top left panel equals 250 µm.

### Characteristics of iPSC selectively inactivated for mutant *BAG3*

Finally, we evaluated if our personalized allele-specific CRISPR-Cas9 approaches could produce autologous cell resources that are suitable for regenerative medicine. Specifically, we determined whether our allele-specific CRISPR strategies altered the characteristics of iPSC or interfered with the capability of differentiating into cardiomyocytes. We selected representative targeted clonal lines for immunostaining and real-time PCR (qRT-PCR) assays. As shown by immunostaining assays (S. Fig. 6) and quantitative qRT-PCR (data not shown), both iPSC lines from a non-disease control and two study subjects showed qualitatively similar characteristics of iPSC (i.e., OCT4, SSEA4, NANOG, TRA1-40). Furthermore, the inactivation of mutant *BAG3* did not alter the features of iPSC, suggesting that one functional copy of *BAG3* is sufficient to maintain the characteristics of iPSC. In addition, the inactivation of mutant *BAG3* did not interfere with the differentiation capability of iPSC. For example, expression of CTNT, indicative of muscle cells, was qualitatively indistinguishable between cardiomyocytes differentiated from unmodified patient-derived iPSC and targeted iPSC carrying knock-out mutation on the mutant *BAG3* (Fig 5).

## Discussion

Due to limitations in base editing, prime editing, and CRISPR-Cas9 approaches that are aimed at correcting disease-causing mutations, we reasoned that inactivation of mutant *BAG3* may represent an alternative approach with an increased feasibility. Since dominant mutations cause numerous diseases, allele-specific CRISPR-Cas strategies that selectively inactivate the disease allele are broadly applicable to address human diseases [29, 31]. Taking advantage of the strong requirement of NGG PAM sequence for SpCas9 [30], we previously developed allele-specific CRISPR-Cas9 approaches, capitalizing on genetic variations that generate or eliminate the NGG PAM sequence, taking Huntington’s disease as an example [29]. As expected, our PAM-altering SNP-based CRISPR-cas9 strategies showed high levels of allele specificity [27-29], supporting its utility in other dominant disorders. In this study, we developed personalized allele-specific CRISPR-Cas9 strategies for MFM6 using PAM-altering SNPs, and showed their complete allele specificities. In addition, allele-specific CRISPR-Cas9 strategies for MFM6 using the disease-causing mutation as the PAM-proximal SNP were highly selective. Furthermore, the allele-specific CRISPR-Cas9 strategy did not interfere with the iPSC characteristics or the differentiation capacity into muscle cells. Together, our data demonstrate 1) the feasibility of identifying the mutant *BAG3* gene in subjects with *do novo* mutations, 2) high levels of allele specificity of individualized allele-specific CRISPR-Cas9 strategies for MFM6, and 3) the utility in producing cell resources for regenerative medicine for MFM6, offering novel routes for alternative therapeutic development.

Overall, our allele-specific strategies for our study participants did not inactivate the normal *BAG3*, representing one of the advantages. Although the TP-CRISPR strategy for subject #3 utilized one non-allele-specific gRNA due to the lack of a mutant-specific PAM site in the region (Fig. 2A), all edited alleles were mutant *BAG3*. Although this strategy is expected to generate small indels in the upstream region of the normal *BAG3* gene in subject #3, we do not expect that such small indels in the upstream of normal *BAG3* negatively impact the transcription. If using two allele-specific sites is critically important, TP-CRISPR strategies can be further optimized by targeting allele-specific sites generated by PPS. Also, PAS that is far from the TSS can be used to target the genomic region that does not impact the transcription of a gene of interest, as the deletion of a fairly large genomic region is feasible [29], representing another strength.

Despite their promises and strengths, our allele-specific CRISPR-Cas9 strategies to knock out the mutant *BAG3* are limited for various reasons. One potential limitation is that reduced *BAG3* levels may contribute to the DCM, and therefore, we speculate that the ultimate treatment strategy would be gene correction approaches. However, considering 1) the outcomes of DCM are much milder compared to MFM6 and 2) mutant-specific inactivation of *BAG3* at specific developmental stage may not entail DCM, MFM6 patients may get significant benefits from our allele-specific CRISPR strategies while effective gene correction treatments are being developed. In addition, our patient-derived cells treated with allele-specific CRISPR-cas9 strategies did not alter the characteristics of iPSC or their ability to differentiate into neurons (data not shown) and cardiomyocytes, illustrating their utilities in the regenerative medicine setting.

The field has begun to witness clinical applications in the clinic [35-37]. The successful application of allele-specific CRISPR-Cas9 strategies in patient-derived cells not only provides a foundation for targeted therapies in MFM6 but also underscores its utility in investigating underlying disease mechanisms. Taken together, our data, representing the first demonstration of the feasibility of allele-specific CRISPR-Cas9 approaches in patient-derived cells, offer new avenues toward the development of rational treatments for MFM6.

## Methods

### Study approval

Patient consents and the overall study were reviewed and approved by the ethics committee/IRB of Mass General Brigham.

### TOPO cloning and sequencing to determine the phase of *BAG3* mutation in the affected individuals

In order to determine whether the disease-causing *de novo* mutations have occurred on the paternal or maternal chromosome in our study subjects, we performed haplotype phasing analysis, taking advantage of a neighboring heterozygous site. We performed a series of sequencing analyses focusing on a genomic region, encompassing rs121918312 (i.e., p.Pro209Leu mutation) in order to identify a neighbor site with heterozygous genotype in our two affected subjects (i.e., #3 and #6). Among various sites, we used rs196330, which is located approximately 4 Kb downstream of the disease-causing mutation, for subsequent phasing through cloning and sequencing. Briefly, genomic DNA from our probands was amplified by Ex Taq DNA polymerase using specific primers (5’-GCCTCTGACTGCTCATCCTC-3’ and 5’-ATTTCAGGGAGCGGGTTC-3’), and PCR amplicon was cloned into a plasmid using Zero Blunt TOPO PCR Cloning Kit (Thermo Fisher). Subsequently, we performed Sanger sequencing to determine alleles at rs121918312 and rs196330 to infer the phase of the mutation.

### Sanger sequencing to identify PAM-altering SNPs

We performed a series of Sanger sequencing analyses to identify mutant allele-specific NGG PAM sites in our probands. Genomic DNA samples were isolated from our study subjects by DNeasy Blood & Tissue kit (Qiagen, 69506) and amplified by Ex Taq DNA polymerase (Takara). PCR products were purified by QIAquick gel expression kit (Qiagen) and subjected to Sanger sequencing. We focused on identifying NGG PAM site on the paternal and maternal chromosomes in subject #3 and #6, respectively. The following primer sets were used to amplify each region: chr10:121429865 - 121430523, CATCCCCCAGCATCTCATAG and CTCAAGCAGGTGTGAAGCTG; chr10:121430502 - 121431168, TTCAGCTTCACACCTGCTTG and CACTGGGCTGCTCTGAAGAT; chr10:121431093 - 121431793, GAGCGCTTCCTGTGTGCT and GTCAGAGGCAGCTGGAGACT; chr10:121431714 - 121432391, AAGCCAGGGGAGTCATTTGT and AATGATCAGCCCAGCAAAAG; chr10:121432380 - 121433057, GGGCTGATCATTGTGCATT and TGTCCTGCTGCTAAGCTGTG; chr10:121433000 - 121433686, AGGTTCCTGGAAGAGGATGTC and TTAGAGAGGGTGTTCCACAGC; and chr10:121433608 - 121434294, TCAGTCTTCCCTCAGCCTTT and TGGCCAGGCTAGTCTTGAAC. These procedures identified one and two mutant-specific PAM-altering SNPs in subject #3 (rs3847489) and #6 (rs196290 and rs11199061), respectively.

### Cloning of gRNAs

gRNAs for TP-CRISPR strategies (subject #3, GGTCCTCGGCTATCATATAT and CAGACCCTACTTCCATAGCT; subject #6, AGGTGACATTTTAGATCTTT and GACTTTGCAAGGTGCTGTCT) were cloned into lentiCRISPR v2 vector (Addgene, 52961). For NMD-CRISPR, plasmids for Cas9 (PX551; http://n2t.net/addgene:60957) and gRNA (PX552; http://n2t.net/addgene:60958) were obtained from Addgene. To express SpCas9 in iPSC lines, the pMecp2 promoter in the original PX551 plasmid was replaced by an EF-1α core promoter using HindIII/AgeI fragment, generating PX551 EF plasmid. The hSyn promoter for the EGFP marker was also replaced by an EF-1α core promoter using ApaI/KpnI fragment (PX552 EFS plasmid). Cloning of the test gRNAs into the PX552 EFS plasmid was performed according to a recommended protocol (https://media.addgene.org/data/plasmids/60/60958/60958-attachment_wWVpb-8u9Mzp.pdf). Sequences of oligos to generate plasmids are ACCTGCTCGTGTATCACCAGAA and AACTTCTGGTGATACACGAGCA. The resulting vectors were validated by Sanger sequencing.

### Transfection and MiSeq analysis for TP-CRISPR

iPSC from subjects #3 and #6 were co-transfected with gRNA using human stem cell nucleofector kit 1 (Lonza). Cells were treated with puromycin (0.05 µg/ml) for 48 hr, and then DNA samples were isolated for PCR amplification. To evaluate the levels of allele specificity of our TP-CRISPR approaches, nested PCR was performed. Briefly, the 1st PCR assay was performed using a primer set (subject #3, AAGGCAGGTGAGTTTGCACT and AGGCAGGGTACACGAGAGTG; subject #6, TGGACTCCAAAGCGGATAAG and CTCTGGCTTGCTTCTTCCTG) by Ex Taq DNA polymerase. Subsequently, PCR products were purified by QIAquick PCR purification kit (Qiagen), and subjected to the 2nd PCR amplification (subject #3, CTTCCTTCCGGTGTCAGATG and TACAGGCTCCTGACGCACAG; subject #6, GAGGTATTGACTGGGGTTCG and CTCTGGCTTGCTTCTTCCTG) using the same DNA polymerase. Subsequently, Miseq analyses were performed by the Massachusetts General Hospital CCIB DNA core (Cambridge, MA). MiSeq sequence reads were subjected to quality control; the levels of allele specificity were determined based on the allele at rs3847489 and rs11199061 for subject #3 and #6, respectively.

### Transfection and MiSeq analysis for NMD-CRISPR

Cells were transfected with PX551 EFS plasmid and PX552 EFS plasmid by Lipofectamine Stem Transfection Reagent (Invitrogen) according to the manufacturer’s instructions. After 72 hours, cells were harvested and analyzed subsequently. Genomic DNA was isolated from cells using DNeasy Blood & Tissue Kit (Qiagen). Total RNA was extracted using RNeasy Plus Mini Kits (Qiagen). The quality and quantity of RNA were determined using a NanoDrop spectrophotometer (Thermo Scientific). cDNA was synthesized from 50 ng of total RNA using SuperScript IV Reverse Transcriptase (Invitrogen). PCR reactions were performed using Q5 High-Fidelity DNA Polymerase (NEB). Upon ligation of Illumina adaptors and a unique identifier to the amplicon, paired-end sequencing (2x150 bp) was performed by the Illumina MiSeq platform. Deep-sequencing of PCR amplicons was performed by the DNA Core Facility at Massachusetts General Hospital (https://dnacore.mgh.harvard.edu/new-cgi-bin/site/pages/index.jsp). Sequence reads that could not be mapped were removed as part of quality control. Primers for PCR amplification for MiSeq analysis are; rs121918312 For1, CTCTGACTGCTCATCCTCAT and rs121918312 Rev1, TGGGTAGTGCGTCTTCTG for DNA MiSeq; rs121918312 For2, ACCAGGCTACATTCCCATT and rs121918312 Rev2, GGTTTAGAATCCACCTCTTTGC for RNA MiSeq.

### Establishing clonal lines with mutant BAG3 inactivated

To generate single cell clones from iPSCs, cells were transfected with PX551 EFS and PX552 EFS vectors (containing either empty vector or the test gRNA) using Lipofectamine Stem Transfection Reagent. 72 h after transfection without selection, 1.5×10^5^ cells were seeded in a 60 mm dish with CloneR supplement (STEMCELL Technologies) and then incubated for another two weeks for dilution cloning. Visible colonies were picked and individually maintained in 96-well plates for clonal expansion. Once cells reached approximately 80% confluence, cells were sub-cultured in two 96-well plates, one for maintenance and the other for validation of the on-target modification by Sanger sequencing analysis. Single cells were further validated by MiSeq analysis with mRNA.

### Immunoblot analysis for BAG3 protein

Cells were washed twice with cold PBS and lysed with RIPA buffer (Invitrogen) containing protease inhibitor (Roche). Cell debris was removed by centrifugation, and the supernatant was collected. Protein concentration was determined by BCA assays (Thermo Scientific), and samples were denatured by 2X SDS buffer (Invitrogen) with a reducing agent (Invitrogen) for 2 minutes at 80°C. Fifteen micrograms of whole cell lysate was resolved on 4-20% gel (Invitrogen). Transferred membranes were probed by BAG3 antibody (Proteintech, #10599-1-AP) and Actin (Sigma-Aldrich, # A4700).

### Differentiation into cardiomyocytes

For differentiation into cardiomyocytes, iPSCs (clonal lines #3-2 and #6-4) were maintained in mTeSR plus (STEMCELL Technologies) on Matrigel (Corning)-coated 24-well plates. When the confluency of the cell reached approximately 90%, differentiation proceeded according to the manufacturer’s protocol for the cardiomyocyte differentiation kit (STEMCELL Technologies, #05010).

### Immunocytochemistry

Cells were fixed in 4% paraformaldehyde (Thermo Scientific) for 10 min at room temperature (RT) and washed with Dulbecco’s phosphate-buffered saline (DPBS). For permeabilization, 0.5% Triton X-100 (Sigma-Aldrich) in DPBS was added to the fixed cells for 10 min, and a blocking buffer with bovine serum albumin (Sigma-Aldrich) was added into the fixed cells for 1 hour. Cells were rinsed with DPBS and incubated in primary antibody (1 hour) for OCT4 (Cell Signaling TECHNOLOGY, #9656), NANOG (Cell Signaling TECHNOLOGY, #9656), and CTNT (Invitrogen, #MA5-12960). After washing (0.2% Tween 20), cells were incubated in secondary antibody (rabbit IgG, Invitrogen, #A-21206; mouse IgG, Invitrogen, #A-11003; 1 hour at RT) and washed with 0.2% Tween 20 in DPBS. For nuclear acid staining, cells were incubated in 4’-6-diamidino-2-phenylindole (DAPI; Thermo Scientific, #62248; 5 min) and washed with 0.2% Tween 20.

### Genomic coordinate

Genomic coordinates were based on GRCh37/hg19.

## Data Availability

All data produced in the present work are contained in the manuscript.

## Acknowledgments

This study would not be possible without the vital contribution of the research participants and their families. Supported by the Alexander’s Way Research Fund and NINDS (NS105709 and NS119471).

## Author contributions

JML designed the study. JWS, KHK, YL and DEC performed experiments. JWS, KHK and YL analyzed data. JML wrote the manuscript with the help of JWS, KHK, and YL.

## Supplementary Material

**Supplementary Figure 1.**
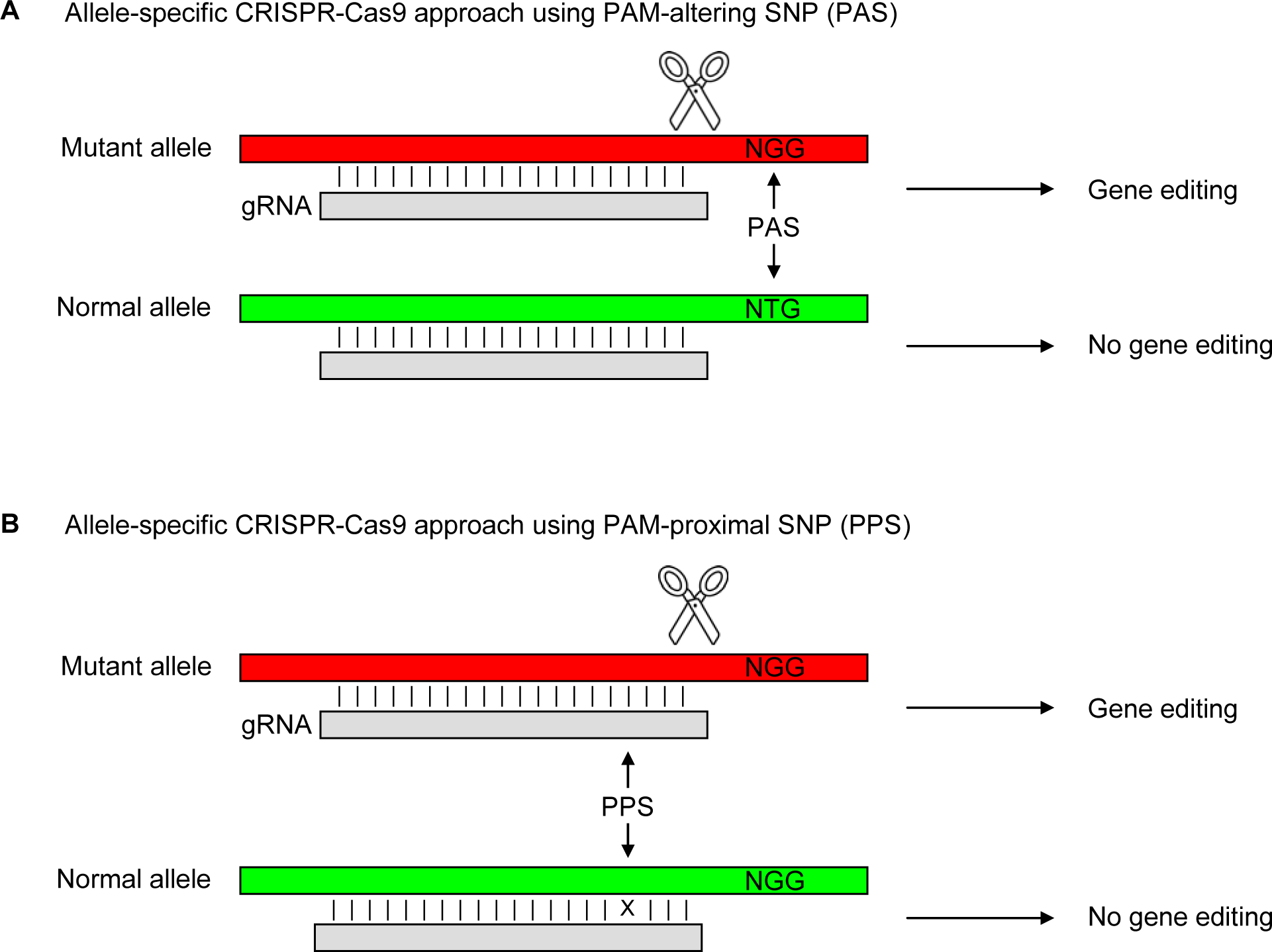
Allele-specific CRISPR-Cas9 approaches utilizing PAS and PPS. A) A PAM-altering SNP may generate the NGG PAM site selectively on the mutant allele (red) in a given individual. The same gRNA relying on this PAM site is, therefore, expected to edit the mutant allele, leaving the normal allele (green) intact. B) If a SNP flanked by a nearby PAM site, that also serves as a target of allele-specific CRISPR-Cas9. The same gRNA designed for the mutant allele (red) may interact less efficiently with the normal allele due to a mismatch (X), permitting allele-specific gene editing. Scissors represent Cas9. Each vertical line represents a match between the target and gRNA.

**Supplementary Figure 2.**
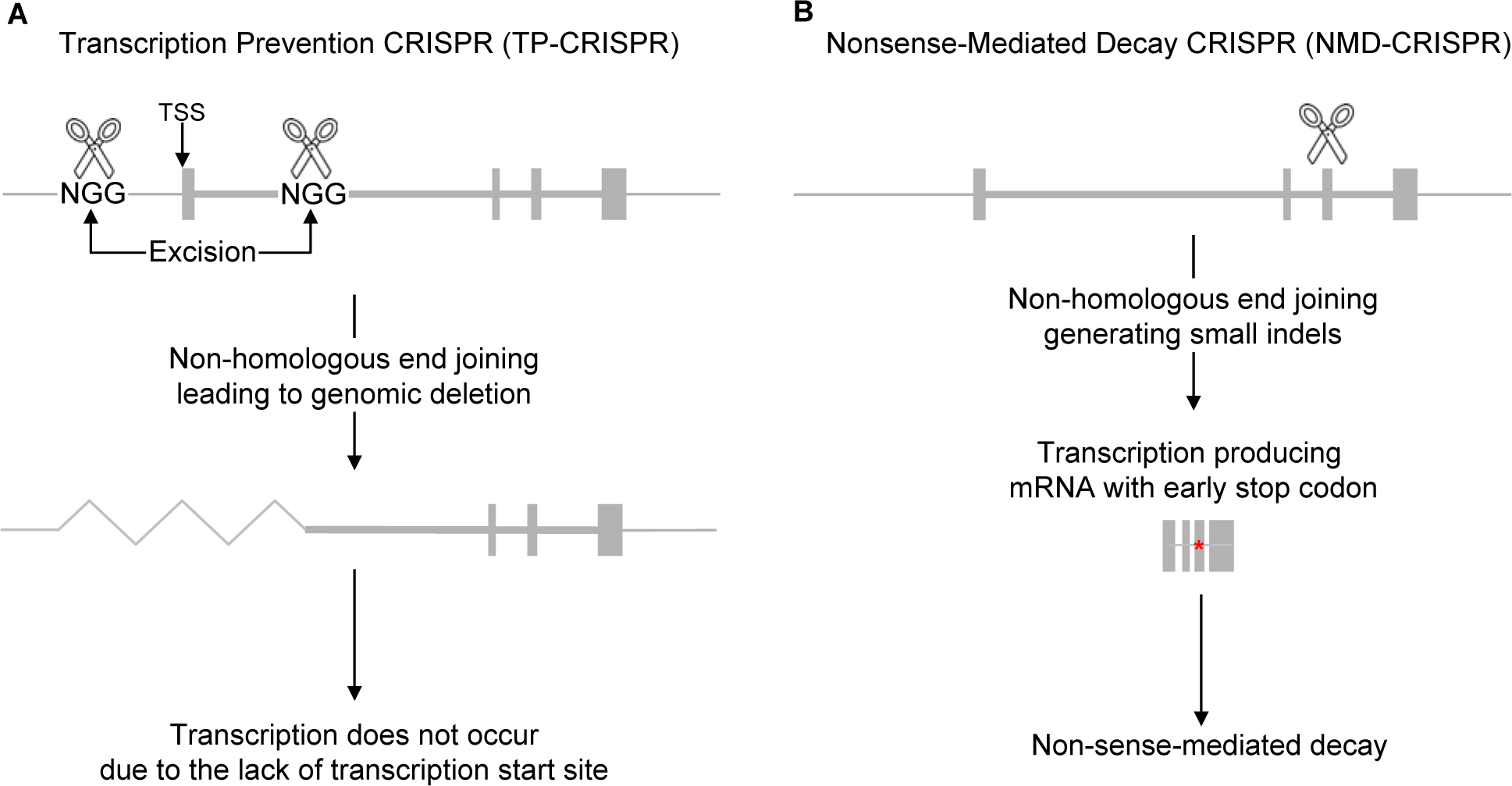
Two modes of CRISPR-Cas9 approaches to inactive a target gene. A) Simultaneous use of two gRNAs, one and the other respectively targeting upstream and the downstream of the transcription start site (TSS), is expected to result in genomic excision. Therefore, two gRNA CRISPR-Cas9 approaches designed to excise the TSS (i.e., Transcription Prevention-CRISPR approach) are anticipated to prevent the transcription from the edited gene. B) Alternatively, a CRISPR-Cas9 strategy designed to edit an exon is likely to generate small indels and, therefore, premature stop codon (a red asterisk), leading to nonsense-mediated decay of the mRNA from the edited gene. For illustration purposes, *the BAG3* gene is depicted. Scissors represent Cas9.

**Supplementary Figure 3.**
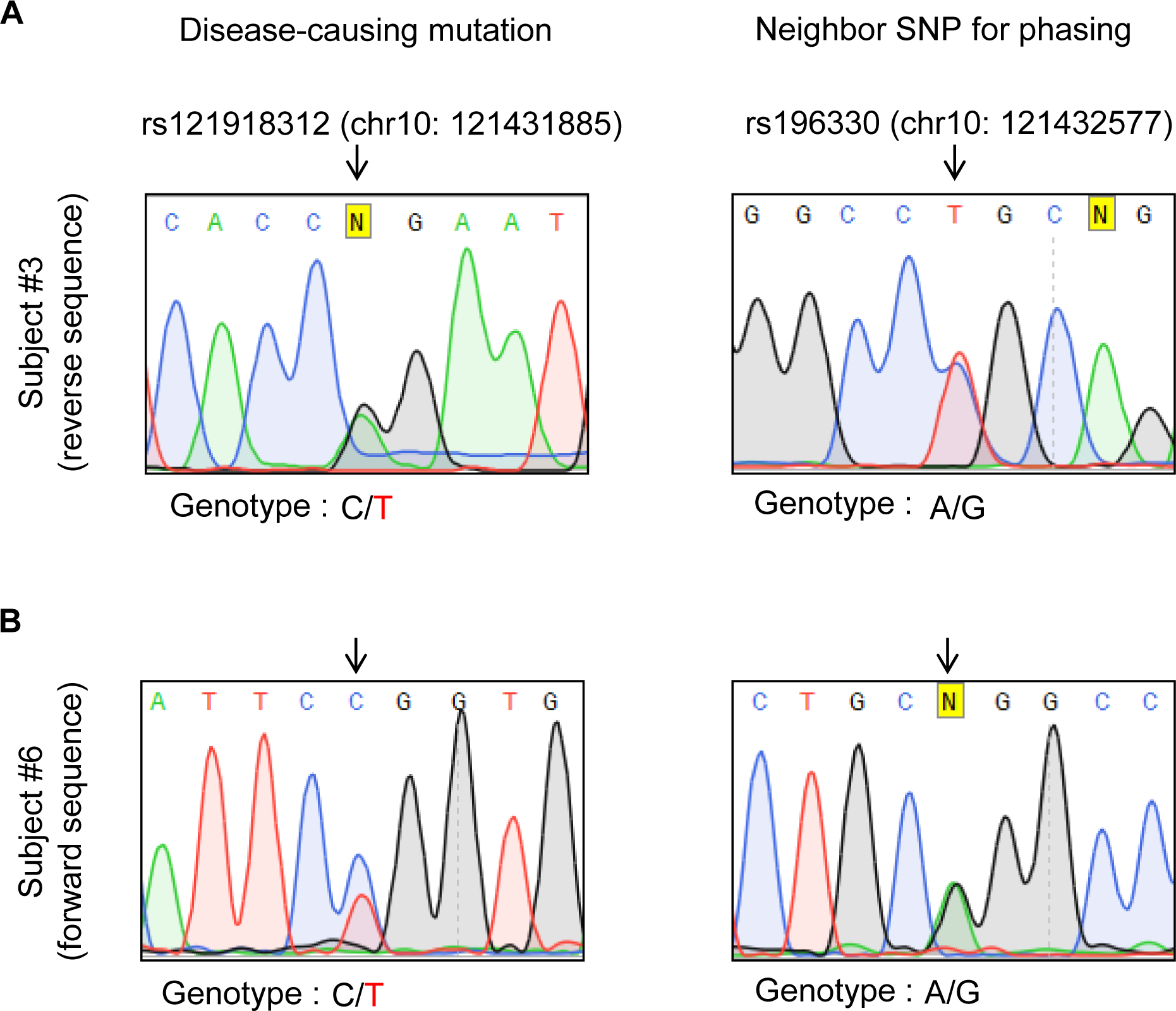
Sanger sequencing results for the disease-causing mutation and a neighbor SNP. A) Sanger sequencing results of subject #3 at rs121918312 (disease-causing mutation) and rs196330 (a neighbor SNP for haplotype inference) are shown. Reverse sequencing results are displayed. B) Sanger sequencing results of subject #6 at rs121918312 and rs196330 are shown. Forward sequencing results are displayed.

**Supplementary Figure 4.**
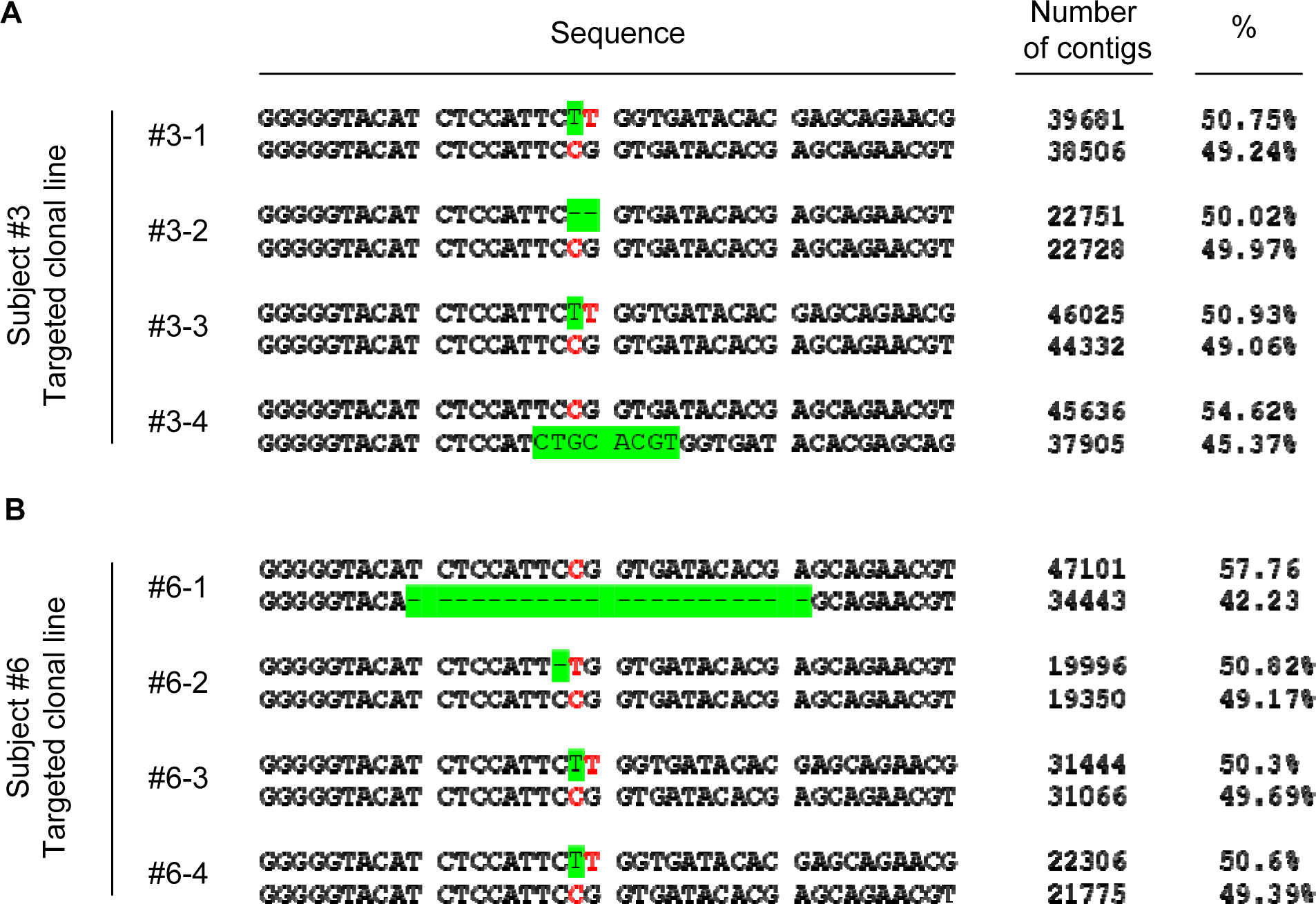
Characterization of targeted clonal patient-derived iPSCs. Following the application of allele-specific NMD-CRISPR, four independent clonal lines with mutant *BAG3* inactivated were established for subjects #3 (A) and #6 (B). The number of contigs and % values represent alleles in the MiSeq sequencing analysis for each clonal line. Green highlights with letters and “-” represent addition and deletion, respectively. No DNA modification has occurred on the normal allele, which carries the C allele.

**Supplementary Figure 5.**
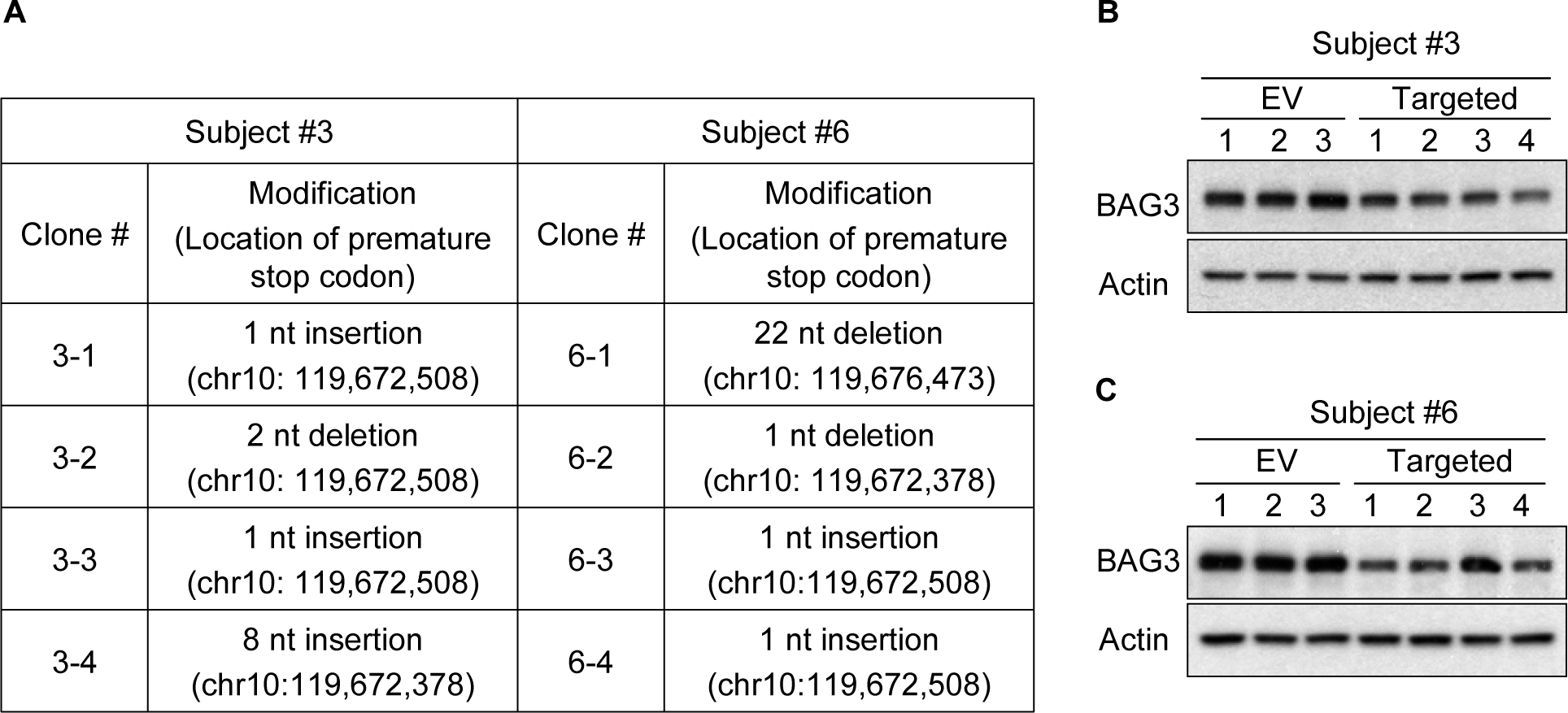
Effects of allele-specific NMD-CRISPR. A) Details of DNA modification of each edited clonal line are summarized. The locations of newly generated premature stop codons are also provided in parentheses. Immunoblot analyses were performed to determine the levels of total BAG3 protein in non-targeted patient-derived iPSC (EV, empty vector) and clonal lines targeted by allele-specific NMD-CRISPR (targeted) from subjects #3 (B) and #6 (C).

**Supplementary Figure 6.**
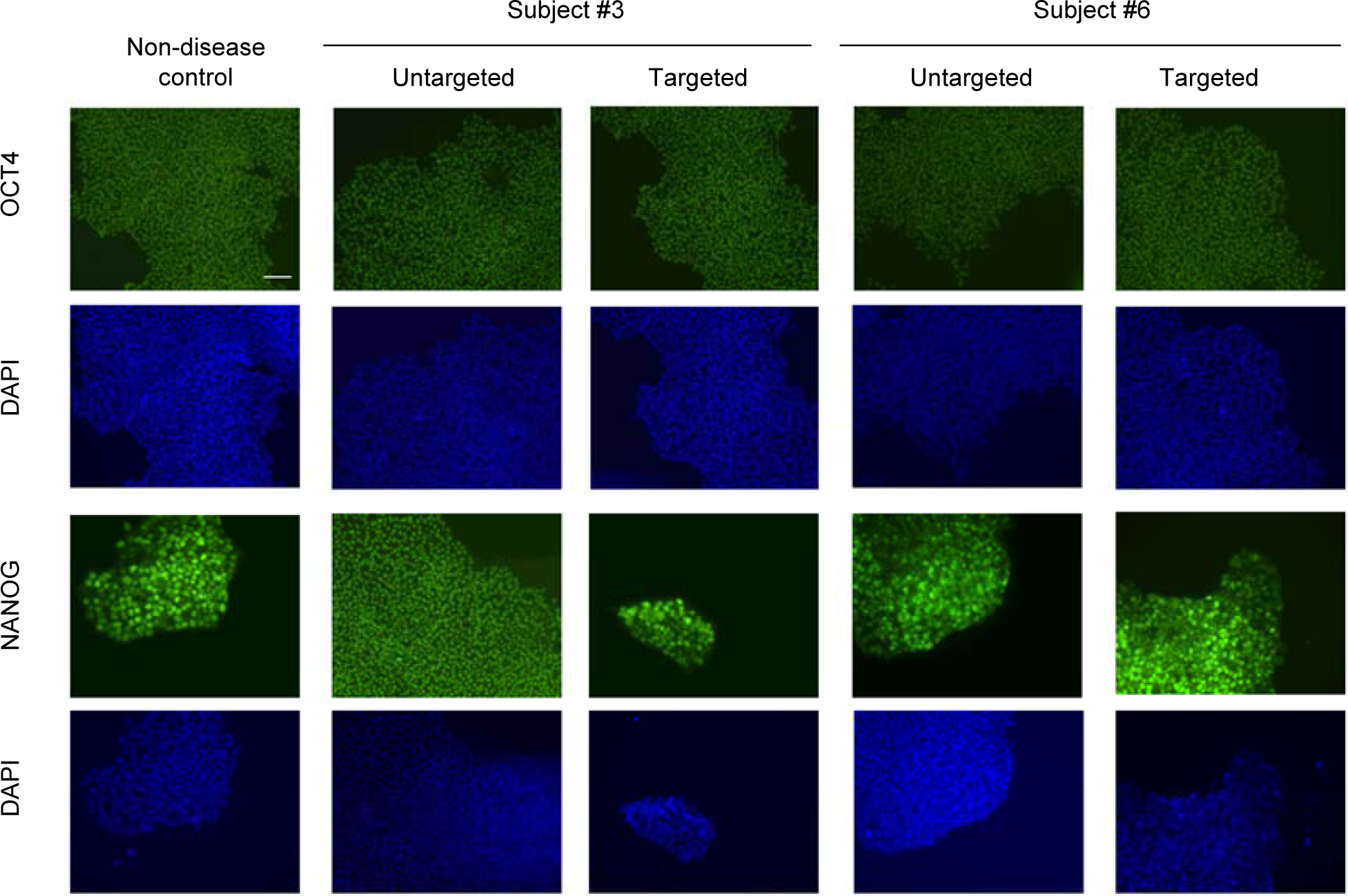
Characterization of iPSC derived from MFM6 patients. Immunocytochemistry of OCT4 and NANOG was performed to evaluate the pluripotency of non-disease control, untargeted, and targeted clonal lines for subjects #3 (clonal line #3-2) and #6 (clonal line #6-4). DAPI staining shows nuclei. The scale bar equals 250 µm.

